# Light Exposure Differs by Gender in the US: Women Have Less Bright Light Exposure than Men

**DOI:** 10.1101/2024.04.28.24306495

**Authors:** Danielle A. Wallace

## Abstract

Light is a salient environmental exposure, serving as the primary entraining cue for the circadian system and having other, non-circadian, effects on health. Gender differences in light exposure patterns could contribute to gender differences in health outcomes and would have important implications for sleep and circadian research. Gender differences in real-world light exposure (measured over a week with wrist-worn ActiGraph GT3X+ devices) were investigated in cross-sectional data from the 2011-2014 National Health and Nutrition Examination Survey (NHANES). Measures of time above light threshold (TALT), individual photoperiod (IP), first and last timing of light (FTL and LTL, respectively), and mean light timing revised (MLiTR) at different light intensity thresholds were derived. Gender differences in light exposure were tested using two-sample t-tests, Watson’s two-sample test of homogeneity, and linear regression models. Exploratory analyses to investigate work and physical activity-related factors in relation to bright light exposure were also conducted. A total of 11,318 NHANES participants (age range: 3-80+, 52.2% women) with 6 days of valid actigraphy and light data were included in the analysis. The findings suggest that for every 60 minutes of bright light (≥1,000 lux) that men receive, women receive 39.6 minutes. Men spend approximately 52% more time in bright light than women and this gender difference begins in childhood. The IP of bright light exposure is also longer for men, with earlier first and later last timing of bright light exposure compared to women. These gender differences were robust across ages and between race and ethnicity groups. While further research is needed, these gender differences in light exposure may be due to gender differences in indoor vs. outdoor activities. Future studies of gender differences in response to light exposure should consider light exposure history in study design and analysis. The results of this study may inform future health disparities research and support the importance of the study of light as an important environmental exposure and component of the human exposome.

## INTRODUCTION

Light is a salient environmental exposure, serving as the primary entraining cue for the circadian system and having other, non-circadian, effects on health. While electric light is now common in the built environment, it wasn’t until the mid-1920’s that approximately half of U.S. households had access to indoor electric lighting^1^. Nearly one hundred years later, electric lighting is widespread but there is surprisingly little objective evidence characterizing personal light exposure patterns in the general population.

Just as air and water are environmental elements important for public health, light is an environmental exposure worthy of scientific measurement and attention. However, unlike monitoring of other environmental exposures and pollutants, light exposure has not been well-integrated into biomonitoring efforts and environmental health research. Additionally, most prior studies of light exposure have been conducted in experimental or clinical settings or rely on satellite imagery to derive outdoor light exposure measures, and therefore do not capture indoor light exposure or personal behavior. Because light may be an important contributor to health and health disparities, it is in the interest of public health to understand real-world light exposure patterns.

Light exposure is implicated in numerous health conditions, such as vision and refractive error, sleep disorders, and mental health. Interestingly, some light-related health conditions also differ by sex and/or gender; for example, men have a higher prevalence of cutaneous melanoma^2^ and females show faster myopia progression^3^, higher prevalence of autoimmune disease^4^, and higher prevalence of insomnia^5^. These differences may be due to biology, or they may be due to environmental, sociocultural, and/or behavioral differences. For example, women have shorter intrinsic circadian periods (τ) and go to bed earlier and wake up earlier^6^ than men. It is also possible that gender and sex differences in light exposure patterns could contribute to gender and sex differences in health outcomes. Examining whether real-world light exposure differs by sex or gender could advance our understanding of sleep and circadian population health and has important implications for experimental light intervention research.

Therefore, to investigate whether gender differences in light exposure exist, we derive measures which reflect multiple dimensions of light exposure (timing, intensity, duration) using objectively measured personal light exposure from the National Health and Nutrition Examination Survey (NHANES) from 2011-2014. Collected using light-sensing wrist-worn actigraphs^7–11^, this data can be used to understand real-world light exposure patterns in the non-institutionalized U.S. population. We test whether gender differences in light exposure patterns exist and explore whether differences are explained by work activities and/or physical activites. The results of this study may inform future health disparities research and promote the study of light as an important environmental exposure and component of the human exposome^12^.

## METHODS

### Study population and characteristics

This analysis used cross-sectional data from the 2011-2012 and 2013-2014 cycles of the Centers for Disease Control and Prevention (CDC) National Health and Nutrition Examination Survey (NHANES), representative of the non-institutionalized U.S. population. Details on the NHANES study have been described elsewhere and are available at: https://www.cdc.gov/nchs/nhanes/about_nhanes.htm. Briefly, questionnaire data and health-related information were collected during physical examination in a Mobile Examination Center (MEC). Gender was either participant-reported or assumed and noted by the interviewer, with the options: “male”, “female”, “don’t know” or “refuse”. It is acknowledged that this item may not appropriately capture gender identity and that the option labels “male” and “female” may reflect sex rather than gender. In general, gender refers to sociocultural constructs and behaviors while sex refers to biological constructs. However, the terms “women” and “men” are hereafter used throughout the text to maintain consistency with the gender label of the NHANES variable. Body mass index (BMI) was measured during the physical exam. Questionnaires and/or interviews were used to collect data on self-reported race and ethnicity, self-reported work and physical activities, and self-reported time spent outdoors. Race and ethnicity were included as a covariate in some models to attempt to account for the influence of racism and bias on characteristics that might impact light exposure, such as the built environment. Time spent outdoors was only asked for participants aged 20-59. Further data processing details are described in **Supplemental Materials**. This analysis included participants with valid actigraphy and light data. The Ethics Review Board of the CDC National Center for Health Statistics approved NHANES. All participants provided informed consent.

### Actigraphy measurement and data pre-processing

Following the physical exam, a subset of NHANES participants wore a wrist-worn GT3X+ ActiGraph device (ActiGraph, Pensacola, FL) for up to 9 days. This device concurrently measures light exposure and triaxial movement in 1-minute epochs. NHANES collected activity and light data from wrist-worn GT3X+ ActiGraph devices (ActiGraph, Pensacola, FL) across 9 days of measurement from a subset of the study sample following the MEC exam. Minute- epoch actigraphy data were downloaded and processed. In addition to existing missingness, if the epoch was flagged for quality by NHANES, if device was predicted to be off-wrist, or if the activity value was <0 then the light and activity values for this epoch were also set to missing. Light and activity measures were then derived from the first 6 valid days of wear, where a valid day was defined as a day with ≤6 hours of missingness and daily activity sum count ≥ 200.

Further details are provided in **Supplemental Material**.

### Creation of light variables

For average light and activity measures, daily summary measures were first created before being averaged across days. To calculate the duration of time spent in different light intensities (time above lux threshold (TALT), **Supplemental Table 1**), a binary variable was first created to indicate whether an epoch’s lux value was below, within, or above a particular threshold (1-9 lux, 10-99 lux, 100-999 lux, ≥1,000 lux). In general, light exposure ≥1,000 lux likely represents outdoor light (sunlight), 100-999 lux may represent indoor or outdoor environments for the GT3X+^13^, and light exposure <100 lux likely represents indoor environments (or outdoor nighttime environments). For the individual photoperiod (IP) and mean light timing revised (MLiTR) variables, first and last daily timing (FTL and LTL, respectively) of light at particular thresholds (≥ 10 lux, ≥ 100 lux, and ≥1,000 lux) were derived by taking the first or last epoch with 3 consecutive epochs (3 minutes) of light exposure at the lux threshold (**Supplemental Table 1**). IP duration (hours) was calculated by taking the daily difference in timing between FTL and LTL at a particular threshold; MLiTR was also calculated as the halfway point between the first and last TALT occurrence (time, Hour:Minute). Further details are provided in **Supplemental Material**.

### Statistical analysis

Data were analyzed with 4-year (2011-2012, 2013-2014) combined MEC sample weights (https://wwwn.cdc.gov/nchs/nhanes/tutorials/module3.aspx) using the “survey” package^14^ to derive population-based estimates, except for the time-based circular variables. The mean, 95% confidence interval of the mean, and/or standard deviation (SD) of light variables were calculated as summary measures. The “circular”^15^ R package was used for circular statistics calculations. Plots of both unweighted and weighted data (using sample population weights) are shown. For variables related to timing (FTL, LTL, MLiTR), the circular (vector-based) mean and SD were calculated. Population-weighted two-sample t-tests and Watson’s two-sample test of homogeneity (for circular variables) were used to test for gender differences in light measures. A p-value <0.05 was considered statistically significant. All analyses were performed in R version 4.1.1

## RESULTS

There were 11,318 NHANES participants with valid light and actigraphy data included in this analysis (**Figure 1**). The average age of participants was 38 years old (age range: 3-80+ years), with 52% women (**Table 1**). By race and ethnicity groups, approximately 4.6% of the sample self-reported belonging to the non-Hispanic (NH) Asian group, 11.7% to the NH Black group, 64.4% to the NH White group, 10.1% to the Mexican American group, 6.2% to the Other Hispanic group, and 3.0% to the Other or Multiracial group. The average duration of time spent in bright light (TALT_1000_) was 1.09 hours daily (95%CI: 0.96, 1.21 hours), moderate-bright light (TALT_100-1000_) was 2.22 hours daily (95%CI: 2.11, 2.33 hours), dim-moderate light (TALT_10-100_) was 4.09 hours daily (95%CI: 3.96, 4.21 hours), and dim light (TALT_1-10_) was 1.42 hours daily (95%CI: 1.39, 1.44 hours) among included participants.

**Figure 1.**
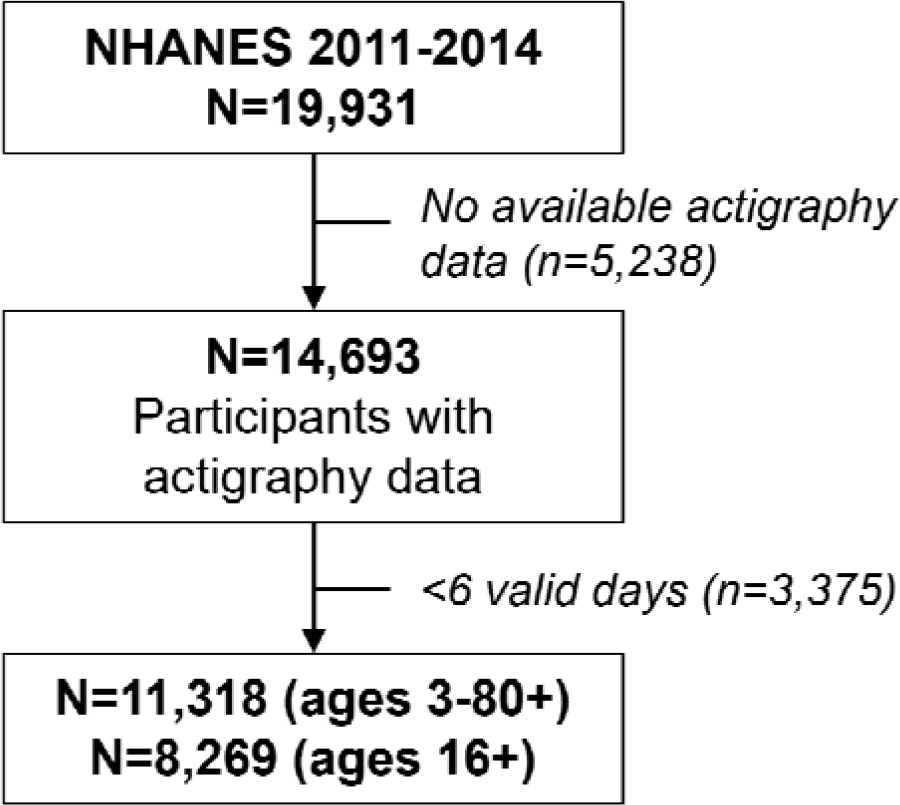
Flow chart showing the sample sizes of participants included in the analyses.

**Table 1.** Demographic characteristics of the 2011-2014 NHANES participants (ages 3-80+ years) included in the analysis.

|  | **All**<br>(n=11,318) | **Men**<br>(n=5,513) | **Women**<br>(n=5,805) |
| --- | --- | --- | --- |
| **Age, years** (mean (SD)) | 38.1 (21.8) | 40.5 (21.4) | 42.3 (21.6) |
| **BMI** (mean (SD)) | 26.9 (7.6) | 27.2 (6.9) | 27.8 (8.0) |
| **Gender** (n (% women)) | 5,513 (52.2) | - | - |
| **Race/ethnic group** (n (%)) |  |  |  |
| *NH Asian* | 1,239 (4.59) | 584 (4.36) | 655 (4.79) |
| *NH Black* | 2,780 (11.66) | 1,357 (11.15) | 1,423 (12.13) |
| *NH White* | 4,012 (64.41) | 1,963 (64.03) | 2,049 (64.76) |
| *Mexican American* | 1,749 (10.11) | 880 (11.01) | 869 (9.29) |
| *Other Hispanic* | 1,108 (6.19) | 514 (6.11) | 594 (6.26) |
| *Other/Multiracial* | 430 (3.04) | 215 (3.34) | 215 (2.76) |
| **Season of measurement** (n (% May-October)) | 5,718 (54.2) | 2,711 (53.10) | 3,007 (55.23) |
| **Family income below federal poverty level** (n (% yes)) | 2,965 (18.54) | 1,353 (17.02) | 1,612 (19.94) |

*Note: the percentages, means, and standard deviations provided in the table are population-weighted; the sample number reflects raw participant sample numbers*

### Women have less bright light exposure compared to men

Across all ages, men spent approximately 52% more time in bright light compared to women (men TALT_1000_=1.32 (95%CI: 1.16, 1.48 hours); women TALT_1000_=0.87 (95%CI: 0.77, 0.98 hours); **Figure 2**). Therefore, women received 39.6 minutes of bright light for every 60 minutes of bright light exposure that men received. This gender difference emerged in childhood but was greater for participants aged 16 and older, with men having approximately 58% more time in bright light, characteristic of the outdoor environment, compared to women (**Table 2**). The results were similar with (**Supplemental Figure 1**) or without (**Figure 2**) applied sample weights. This effect did not meaningfully differ with or without adjustment for age, race and ethnicity, and season (β=-0.45 for the effect of gender (ref=men) in unadjusted and adjusted models, p<0.001). Women also spent greater time in dim light environments, characteristic of the indoor environment, compared to men (men TALT_10-100_ = 3.85 (95%CI: 3.73, 3.97 hours); women TALT_10-100_ = 4.30 (95%CI: 4.16, 4.45 hours); **Figure 2**), although this difference was most pronounced in participants 16 and older (**Table 2**). The gender gap in bright light existed across all race and ethnicity groups except for the Other or Multiracial group; however, this group had a smaller sample size and may have been underpowered to detect a difference. Differences were most pronounced among participants aged 16 years and older (**Table 3**). The mean difference in TALT_1000_ exposure between men and women aged 16 and older was greatest for the Mexican American group (mean difference=-0.73) followed by the Other Hispanic group (mean difference=-0.57), the NH White group (mean difference=-0.51), and the NH Black group (mean difference=-0.47; **Table 3**).

**Figure 2.**
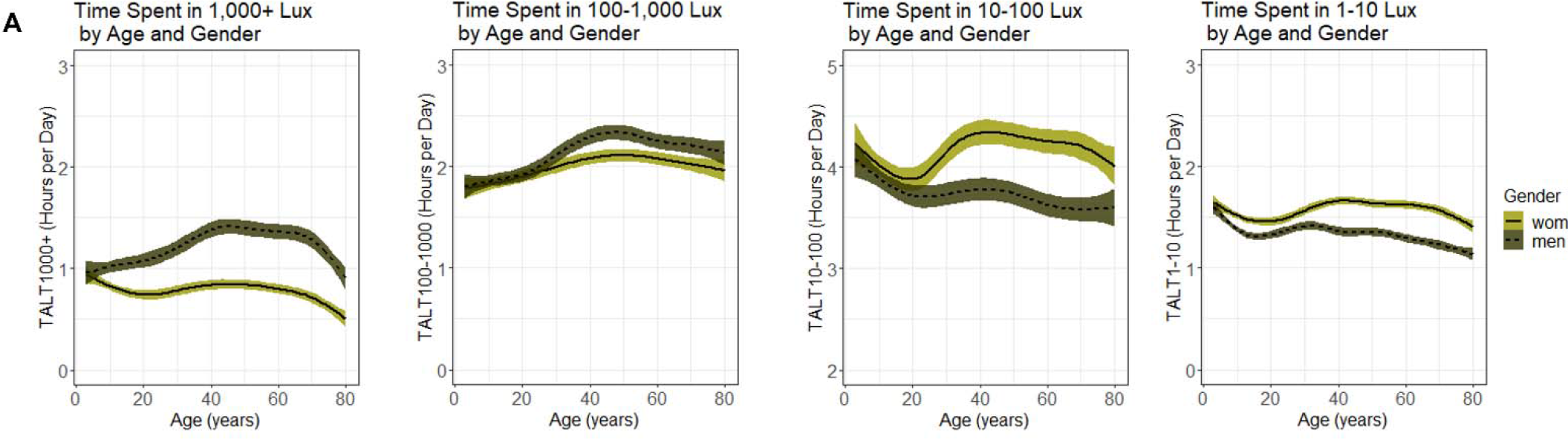
Plots showing the (**A**) average time spent above light threshold (TALT) at different intensities (≥1,000 lux, 100 to <1,000 lux, 10 to <100 lux, and 1 to <10 lux) across ages and by gender (unweighted for sample weights). Dashed lines indicate men and solid lines indicate women. Colored shading around black line (mean) indicates the 95% CI.

**Table 2.**
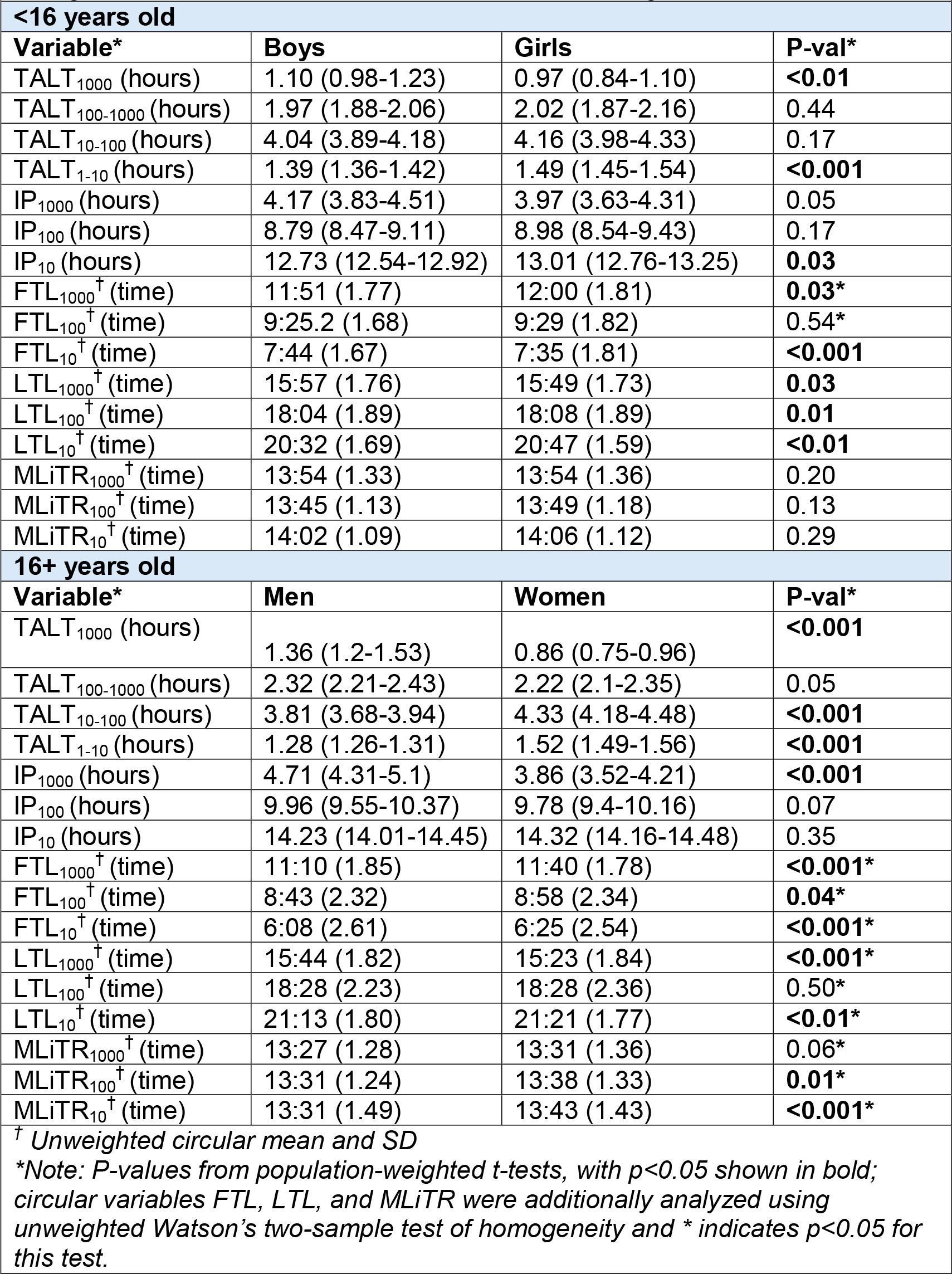
Table comparing light exposure variables with mean and 95%CI or SD by gender in NHANES 2011-2014, stratified by participants <16 years old and ages 16 and older. TALT and IP show population-weighted estimates, while the timing-related variables FTL, LTL, and MLiTR are unweighted.

**Table 3.** Table comparing TALT_1000_ with mean and 95%CI or SD by race and ethnic groups and gender in NHANES 2011-2014, stratified by participants <16 years old and ages 16 and older.

| **<16 years old** |  |  |  |
| --- | --- | --- | --- |
|  | **Boys** | **Girls** | **P-val\*** |
| NH Asian | 0.73 (0.58-0.89) | 0.69 (0.57-0.81) | 0.63 |
| NH Black | 0.95 (0.81-1.08) | 0.68 (0.59-0.77) | **<0.001** |
| NH White | 1.23 (1.04-1.42) | 1.16 (0.97-1.36) | 0.36 |
| Mexican American | 1.04 (0.91-1.18) | 0.77 (0.68-0.87) | **0.01** |
| Other Hispanic | 0.98 (0.85-1.12) | 0.86 (0.71-1.02) | 0.22 |
| Other/Multiracial | 0.92 (0.71-1.13) | 0.90 (0.76-1.04) | 0.84 |
| **16+ years old** |  |  |  |
|  | **Men** | **Women** | **P-val\*** |
| NH Asian | 0.72 (0.59-0.86) | 0.54 (0.44-0.65) | **<0.01** |
| NH Black | 1.21 (1.06-1.37) | 0.74 (0.66-0.82) | **<0.001** |
| NH White | 1.44 (1.23-1.65) | 0.93 (0.80-1.06) | **<0.001** |
| Mexican American | 1.44 (1.26-1.63) | 0.72 (0.63-0.80) | **<0.001** |
| Other Hispanic | 1.25 (1.03-1.48) | 0.69 (0.60-0.78) | **<0.001** |
| Other/Multiracial | 1.15 (0.73-1.58) | 0.90 (0.71-1.09) | 0.15 |
| *Note: P-values for TALT<sub>1000</sub> from population-weighted t-tests stratified by race and ethnicity groups. P<0.05 are shown in bold. |  |  |  |

### The timing of light exposure also differs by gender

In addition to duration of time spent in different light environments, the IP and timing of light also differed by gender. Across all ages, women’s bright light exposure began later and ended earlier in the day compared to men, resulting in a shorter individual photoperiod in bright light. Among participants aged 16 and older, on average, the IP_1000_ for women was approximately 18% shorter than that for men (men IP_1000_=4.71 hours; women IP_1000_=3.86 hours; **Table 2**, **Figure 3**). The first daily timing of bright light exposure for women occurred 30 minutes after that for men (men FTL_1000_=11:10AM; women FTL_1000_=11:40AM; **Table 2**, **Figure 3**) while the last daily timing of bright light exposure occurred approximately 21 minutes earlier (men LTL_1000_=3:44PM; women LTL_1000_=3:23PM; **Table 2**, **Figure 3**). While the IP at lower light intensities did not show gender differences, some of the FTL and LTL measures did show differences, suggestive of later FTL of dim to moderate light and later LTL for dim light for women (**Table 2**).

**Figure 3.**
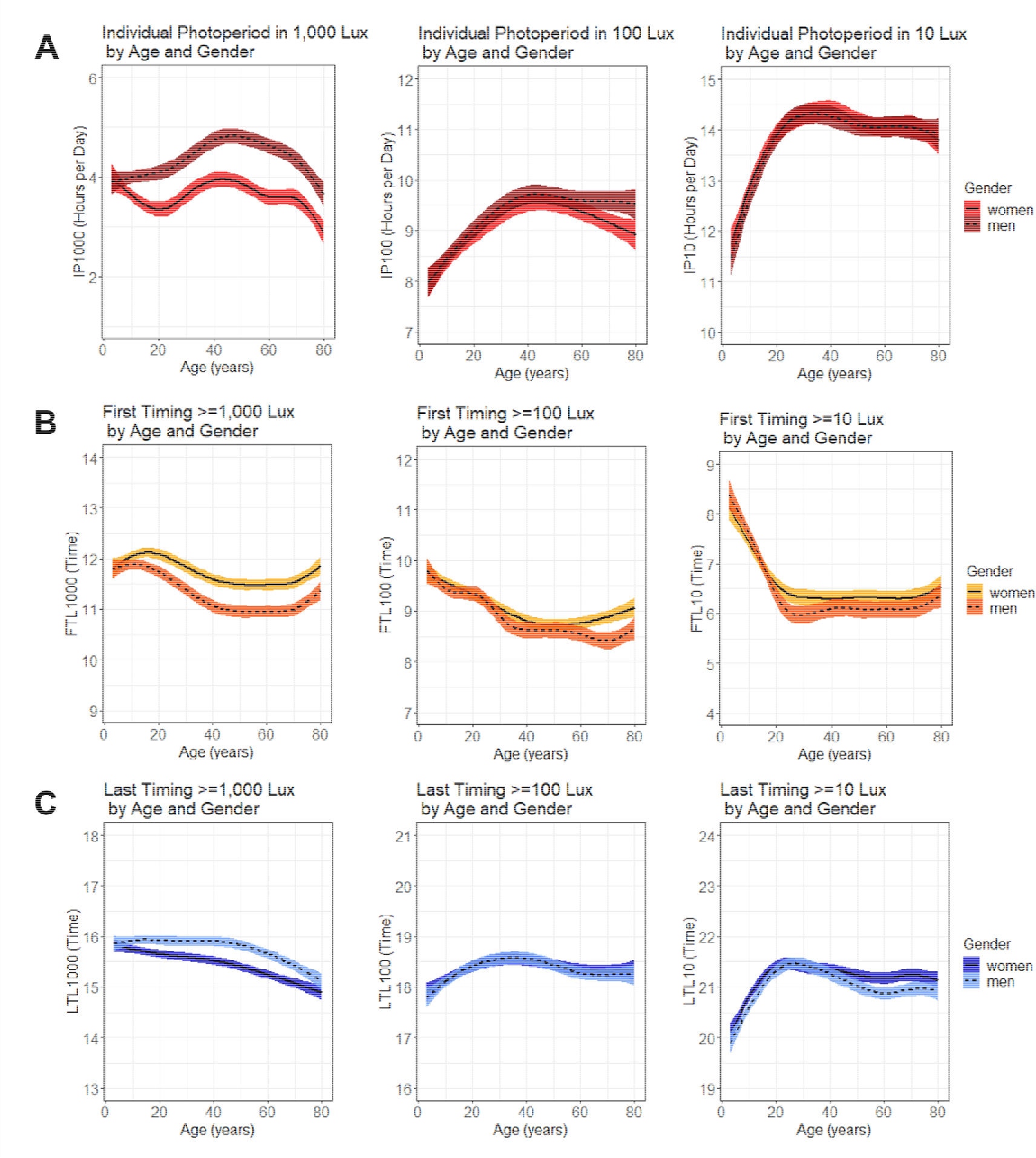
Plots showing the (**A**) average individual photoperiod (IP), (**B**) first timing of light exposure (FTL), and (**C**) last timing of light exposure (LTL) by different light intensity thresholds (≥1,000 lux, ≥100, and ≥10 lux) across ages and by gender (unweighted for sample weights). Dashed lines indicate men and solid lines indicate women. Colored shading around black line (mean) indicates the 95% CI.

### Time spent outdoors may be related to gender differences in light exposure

To better understand the factors that may be related to gender differences in light exposure, self-reported data on time spent outdoors, and work and physical activity were analyzed among participants 16 and older. Men spent 94 more minutes outside 9AM-5PM on workdays and 53 more minutes outside on non-workdays compared to women, for a total difference of >2 hours greater outdoor time (9AM-5PM) for men than women during the week (men total time outdoors=385 minutes (95%CI: 358.7, 411.1); women total time outdoors=242 minutes (95%CI: 230.7, 253.8); **Supplemental Table 2**). While self-reported sedentary activity did not differ by gender, men also reported higher frequency of vigorous and moderate work activities, biking or walking, and vigorous recreational activities compared to women (p<0.05, **Supplemental Table 2**). When these variables were included in an interaction term with gender in separate models testing the association with TALT_1000_, self-reported time outdoors and vigorous or moderate work activities showed a significant interaction with gender, while self-reported walking and/or biking or vigorous recreational activities did not show an interaction (**Supplemental Table 3**).

## DISCUSSION

Women spent less time in bright light compared to men, with men having approximately 52% greater time spent in bright light. Gender differences in light exposure were most pronounced at brighter (1,000+ lux) light intensities. This gender difference existed across most racial and ethnic groups and across age. The average timing for the day’s first bright light was also later for women, occurring approximately 30 minutes after men. While the exact causes of these differences in light exposure are unknown, they likely reflect gender differences in time spent indoors vs. outdoors.

Strikingly, the gender difference in bright light exposure began in childhood, with girls spending less time in bright light than boys. These findings align with prior U.S.-based research, which has reported that girls have 15% lower odds of being taken outside for playtime by their caregivers compared to boys^16^ and spend less time outside on weekdays and weekends^17^.

Possible reasons for these early life disparities in outdoor play may be related to caregiver perception^18^, gendered assumptions or ideas about cleanliness^19^, safety concerns, and/or behavior modeling. There are also gender gaps in outdoor recreation^20^, with women less likely to engage in outdoor physical activity. These differences in behavior may be due to social conditioning and gender norms as well as barriers that discourage girls and women from partaking in outdoor activities. These differences in early-life light exposure could plausibly influence developmental outcomes, such as vision^21^ and immune function^22^; however, research on the developmental impacts of light exposure is in its early stages^23^.

While the data show clear differences in light exposure patterns for women and men, the specific reasons for these differences are difficult to ascertain from the available data. It is unlikely that work or occupational factors are the sole determinant, as gender differences also occurred in childhood and in older age (after general retirement age). When factors related to work activity and physical activity were explored as potential reasons to explain the gender differences in duration of time spent in bright light exposure, the self-reported minutes spent outdoors from 9AM-5PM on workdays and non-workdays, as well as vigorous or moderate work activities, showed interactions with gender in association with TALT_1000_. The finding that women spend more time in dim-moderate light environments (10-100 lux) also likely reflects greater time spent indoors. The 100-1,000 lux range may represent a mixture of indoor and outdoor environments for the GT3X+ device^13^, which is perhaps why men also have greater duration of time in this light level. Greater outdoor time among men may also be related to physical activity.

Prior analyses of NHANES data has reported higher prevalence of physical activity during leisure time among men compared to women^24^. These findings suggest that both work and leisure time in indoor vs. outdoor environments may be an important component in gendered light exposure patterns, although further investigation is needed.

The finding that light exposure is patterned by gender in a nationally-representative study has important implications for sleep and circadian research^25–34^. For example, light exposure history can affect subsequent responses to light exposure, such as the degree of melatonin suppression and phase shift^33,35,36^. Therefore, gender differences in light exposure history could lead to gender differences in responses to later light exposure. In an experiment to test the existence of gender differences in sensitivity to light exposure, the dim light melatonin onset (DLMO) of men (n=27) and women (n=28) aged 18-30 years old was compared in response to a 5-hour light exposure stimulus occurring around the participant’s habitual bedtime. Women were reported to have a stronger suppression of melatonin in response to a 400-lux and 2,000-lux stimulus compared to men, but there were no gender differences in DLMO at lower light intensities^37^. Within women, menstrual phase, progesterone, and estradiol did not appear to influence results; within men, testosterone levels also did not appear to influence results^37^. A separate study in a smaller sample (n=6 men 25-31 years old and n=6 women 22-34 years old) also reported a stronger suppression of melatonin in women compared to men in response to a 2-hour light stimulus (2-4AM, ∼2,000 lux)^38^. These results are in contrast to contradictory studies with smaller sample sizes (n=5 men, n=5 women; 12-1AM, 0- 3,000 lux)^39^ that reported no effect of gender on DLMO light sensitivity. However, the lower light intensities used in one of these studies (n=22 males, n=21 females in 200 lux; n=4 males and n=7 females in 500 lux; 12-1AM)^40^ is close to the 400 lux threshold value shown to have an effect in Vidafar et al^37^. Importantly, however, none of these prior studies examined pre-experiment light exposure history, which may be an important moderator of the effect of light on DLMO and could explain the discrepancies in results if light exposure history is associated with sex or gender. For example, a small experimental crossover study (n=6 females, n=6 males) did not report sex differences in melatonin suppression, perhaps because the study design attempted to maintain similar light exposure history in both males and females in the week prior to testing^41^. Future research on sex or gender differences in the response to light treatment should consider evaluating light exposure history in the study design and analysis.

This study has several strengths and limitations. This analysis used objective light data measured at the individual level in real-world community-based settings from a large sample of participants. The data analyzed in this study is from a nationally-representative sample of the non-institutionalized U.S. population and includes a wide range of ages and race and ethnic groups. However, light exposure in this study was measured from a wrist-worn device (ActiGraph GT3X+), which may not accurately capture light at the eye level. The GT3X device also has a red casing^13^, which may reduce its light measurement sensitivity^42^. Location and exact date data of measurement were not available, so the influence of location and specific time of year on the results was not investigated. There were also limited variables available to explore more nuanced reasons for what may be driving gender differences in light exposure. The findings from this study also reflect the U.S. population and may not be generalizable to other countries.

## CONCLUSION

Light exposure patterns differ by gender in the U.S., with women receiving less bright light and later timing of bright light than men. These gender differences exist across race and ethnicity and age, with gender differences in bright light exposure emerging in childhood. While the causes of these gender differences in light exposure are unclear, they may be related to gender differences in time spent indoors vs. outdoors. These findings have important implications for public health, health disparities, and sleep and circadian research and underscore the importance of considering light as a fundamental component of the environment.

## ACKNOWLEDGEMENTS

### Funding/Support

Supported by funding from the National Institutes of Health (NIH-NHLBI K99HL166700 [to DW]).

### Author Contributions Statement

DAW: conceptualization and study design, methodology, formal analysis, data curation, software, writing and editing of original draft, funding acquisition.

### Access to Data and Data Analysis

The dataset and information regarding study design, measurement, and variables used in this secondary analysis are publicly available online from the CDC’s NHANES website: https://www.cdc.gov/nchs/nhanes/index.htm.

