## Supplemental Materials for "Light Exposure Differs by Gender in the US: Women Have Less Bright Light Exposure than Men"

Danielle A. Wallace^1,2^, MPH, PhD

*^1^Division of Sleep Medicine, Harvard Medical School, Boston MA, USA*

*^2^Division of Sleep and Circadian Disorders, Departments of Medicine and Neurology, Brigham and Women’s Hospital, Boston MA, USA*

**Data availability and preprocessing**

All 2011-2012 and 2013-2014 cycle NHANES data used in this analysis are publicly available from: <https://wwwn.cdc.gov/nchs/nhanes/ContinuousNhanes/Default.aspx?BeginYear=2011> and <https://wwwn.cdc.gov/nchs/nhanes/continuousnhanes/default.aspx?BeginYear=2013>. Light and activity data were collected from a subset of NHANES participants who agreed to wear a wrist-worn GT3X+ ActiGraph device (ActiGraph, Pensacola, FL) for the week following the physical exam. NHANES capped the range of light values 2,500 lux, for a range of 0-2,500 lux. The physical activity monitor minute-epoch actigraphy data for the 2011-2012 and 2013-2014 cycles was downloaded from: <https://wwwn.cdc.gov/Nchs/Nhanes/2011-2012/PAXMIN_G.XPT> and <https://wwwn.cdc.gov/Nchs/Nhanes/2013-2014/PAXMIN_H.XPT>. Light and activity data for a specific epoch were set to missing if any of these NHANES-provided quality control metrics occurred in the epoch: (1) if PAXFLGSM variable had a letter value; (2) if device was predicted to be off-wrist with PAXPREDM=3[^1^](https://sciwheel.com/work/citation?ids=12119620&pre=&suf=&sa=0&dbf=0) or unknown with PAXPRED=4; and (3) if the triaxial activity value was <0. A day was considered invalid if there was more than 6 hours of missing data or if the sum activity count for the day was <200. Participants with <6 valid days of measurement were excluded.

The following NHANES variables, with descriptions, were used in the analysis:

| **NHANES Variable Name** | **Dataset** | **Variable Description** |
| --- | --- | --- |
| RIDAGEYR | Demographics | Age (years; >80 coded as 80) |
| RIAGENDR | Demographics | Gender |
| RIDRETH3 | Demographics | Race and ethnicity categories |
| RIDEXMON | Demographics | Season of measurement |
| INDFMPIR | Demographics | Ratio of family income to poverty |
| BMXBMI | Body measures | Body mass index |
| DED120 | Dermatology | Minutes outdoors 9am - 5pm work day |
| DED125 | Dermatology | Minutes outdoors 9am - 5pm not work day |
| PAQ605 | Physical activity questionnaire | Vigorous work activity |
| PAQ620 | Physical activity questionnaire | Moderate work activity |
| PAQ635 | Physical activity questionnaire | Walk or bicycle |
| PAQ650 | Physical activity questionnaire | Vigorous recreational activities |
| PAQ665 | Physical activity questionnaire | Moderate recreational activities |
| PAQ680 | Physical activity questionnaire | Minutes sedentary activity |
| PAXDAYM | Physical activity monitor | Day by midnight |
| PAXLXSM | Physical activity monitor | White light exposure in lux |
| PAXMTSM | Physical activity monitor | Actigraphy triaxial activity value |
| PAXFLGSM | Physical activity monitor | NHANES actigraphy quality flag |
| PAXPREDM | Physical activity monitor | Actigraphy coding as awake, asleep, non-wear, or unknown |

| ***Supplemental Table 1.*** *Description of the derived light variables.* | | | | |
| --- | --- | --- | --- | --- |
| **Variable** | **Definition** | **Threshold** | **Summary** | **Light Dimension** |
| **First timing of light (FTL)** | The first daily occurrence of 3 consecutive epochs of light exposure at a particular brightness level | Lux thresholds used: ≥10 lux, ≥100 lux, or ≥1,000 lux | Circular mean and standard deviation (across days) | Timing and intensity |
| **Last timing of light (LTL)** | The last daily occurrence of 3 consecutive epochs of light exposure at a particular brightness level | Lux thresholds used: ≥10 lux, ≥100 lux, or ≥1,000 lux | Circular mean and standard deviation (across days) | Timing and intensity |
| **Mean light timing revised (MLiTR)** | The midpoint between the FTL and LTL at a particular brightness level | Lux thresholds used: ≥10 lux, 100 lux, or 1,000 lux | Circular mean and standard deviation (across days) | Timing and intensity |
| **Time above lux threshold (TALT)** | Number of epochs (minutes) where light exposure met or exceeded a particular brightness level | Lux thresholds used: 1-9 lux, 10-99 lux, 100-999 lux, or ≥1,000 lux | Mean and 95% confidence interval (across days) | Intensity and duration |
| **Individual photoperiod (IP)** | Amount of time between first and last daily occurrence of light exposure at a particular brightness level | Lux thresholds used: ≥10 lux, ≥100 lux, or ≥1,000 lux | Mean and 95% confidence interval (across days) | Intensity and duration |

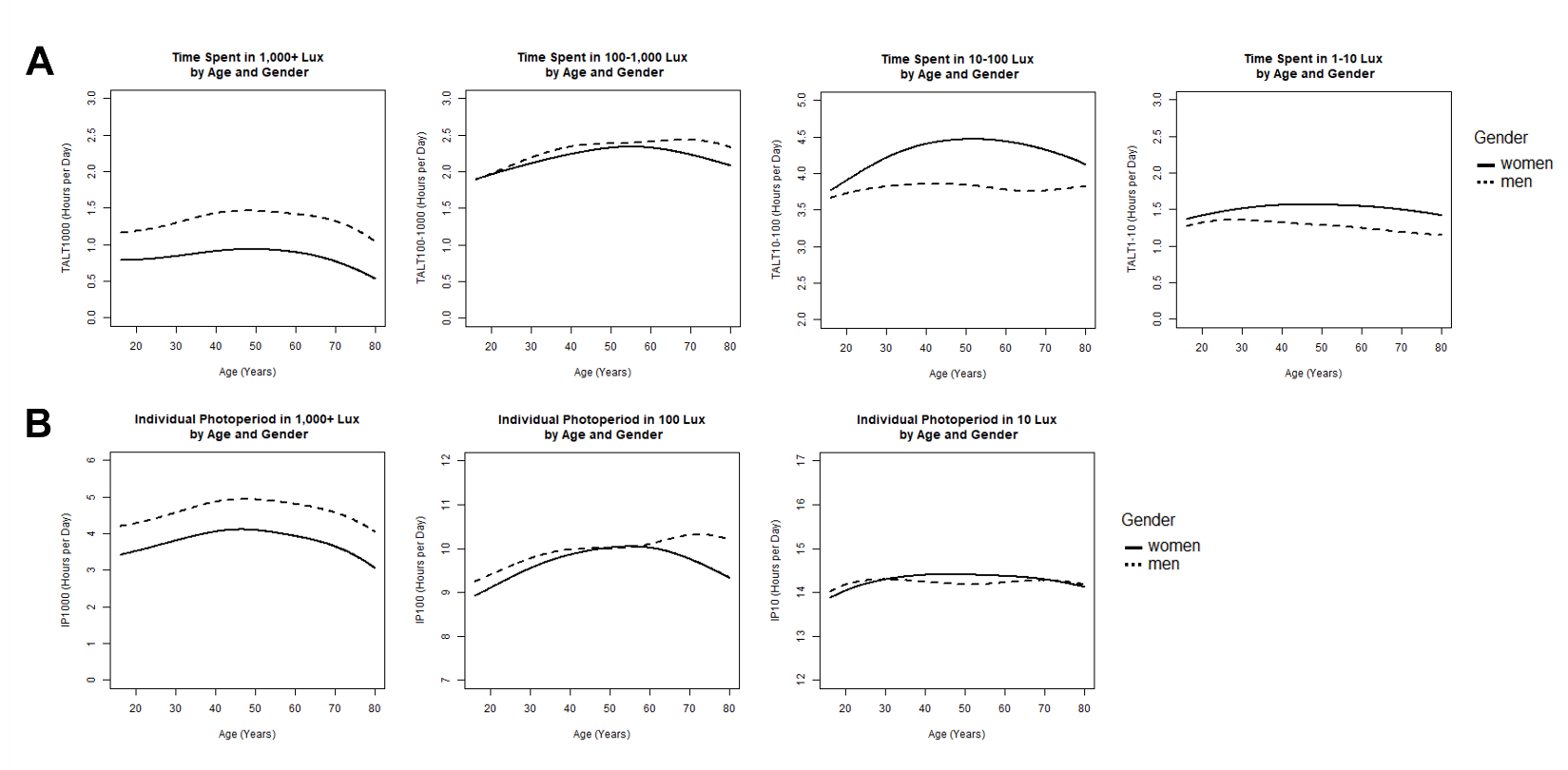

**Supplemental Figure 1**. Plots showing the (**A**) average time spent above light threshold (TALT) and (**B**) average individual photoperiod (IP) at different intensities (≥1,000 lux, 100 to <1,000 lux, 10 to <100 lux, and 1 to <10 lux) across ages and by gender (with applied sample weights). Dashed lines indicate men and solid lines indicate women.

| **Supplemental Table 2**. Table comparing physical activity and occupational variables with mean and 95%CI or SD by gender in NHANES 2011-2014 in participants 16 and older (population-weighted). | | | |
| --- | --- | --- | --- |
| **16+ years old** | | | |
| **Variable** | **Men** | **Women** | **P-val** |
| *Minutes spent outdoors 9am - 5pm on work days | 187.3 (169.9-204.7) | 93.3 (86.1-100.5) | **<0.001** |
| *Minutes spent outdoors 9am - 5pm on non-work days | 194.2 (186.1-202.3) | 141.3 (135.7-147.0) | **<0.001** |
| *Total minutes spent outdoors 9am - 5pm on work and non-work days | 385.1 (358.7-411.4) | 242.3 (230.7-253.8) | **<0.001** |
| Sedentary activity, hours | 6.8 (6.6-7.0) | 6.8 (6.6-7.0) | 0.97 |
| Partake in vigorous work activities, % yes | 26.3% | 10.8% | **<0.001** |
| Partake in moderate work activities, % yes | 39.2% | 31.6% | **<0.001** |
| Use a bicycle or walk to go places (≥10 minutes), % yes | 29.7% | 24.5% | **<0.001** |
| Partake in vigorous recreational activities, % yes | 29.6% | 20.4% | **<0.001** |
| Partake in moderate recreational activities, % yes | 45.0% | 44.7% | 0.85 |
| **Only from participants ages 20-59 years old*  *Note: P-values from population-weighted t-tests and chi-square tests. P<0.05 are shown in bold.* | | | |

| **Supplemental Table 3**. Results from testing interactions of variables with gender on the association with TALT_1000_ in linear regression models adjusted for age, season, and race and ethnicity in NHANES 2011-2014 participants 16 and older (population-weighted). | | |
| --- | --- | --- |
| **Outcome** | **Predictor(s)** | **P-val** |
| TALT_1000_ | Gender  Minutes outside work day  Gender: Minutes outside work day | 0.41  **<0.001**  **<0.001** |
| TALT_1000_ | Gender  Minutes outside non-work day  Gender: Minutes outside non-work day | 0.20  **<0.001**  **<0.01** |
| TALT_1000_ | Gender  Total minutes outside  Gender: Total minutes outside | 0.07  **<0.001**  **<0.001** |
| TALT_1000_ | Gender  Vigorous work activities  Gender: Vigorous work activities | **<0.001**  **<0.001**  **<0.001** |
| TALT_1000_ | Gender  Moderate work activities  Gender: Moderate work activities | **<0.001**  **<0.001**  **<0.001** |
| TALT_1000_ | Gender  Walking or biking  Gender: Walking or biking | **<0.001**  0.34  0.29 |
| TALT_1000_ | Gender  Vigorous recreational activities  Gender: Vigorous recreational activities | **<0.001**  0.58  0.35 |
| *Note: questions regarding minutes spent outside were only asked of participants ages 20-59 years old. P-values <0.05 are shown in bold.* | | |
